# Children with spinal muscular atrophy treated with disease-modifying therapies, do they attain neutral vertical head and trunk control?

**DOI:** 10.1101/2025.02.24.25321598

**Authors:** Tania E. Sakanaka, Penelope B. Butler, Richa Kulshrestha, Tracey Willis, Ian D. Loram

**Author notes:** Correspondence, Address: Department of Life Sciences, Faculty of Science and Engineering, Manchester Metropolitan University; The Dalton Building, Oxford Road, Manchester, M1 5GD, UK.

## Abstract

**Aim:** To investigate whether children with various spinal muscular atrophy (SMA) subtypes treated with disease-modifying therapies (DMT) obtain neutral vertical (NV) head and trunk control, and how this relates to motor function.

**Method:** In this observational cross-sectional study, a convenience sample of 21 children diagnosed with SMA (6y1m (4y), 10 female) was tested with Segmental Assessment of Trunk Control (SATCo) and functional assessments. A linear mixed-effects model (LME) was used for statistical analysis.

**Results:** SATCo scores were measurably different between condition (types of SMA), F=4.96, p<.05, and control type (static, active, and reactive control), F=6.40, p<.05. The interaction condition*control type was also significant, F=3.14, p<.05. No child with SMA1 demonstrated NV head control and there was no relationship between SATCo scores and motor function. Four children with SMA2 and SMA3 demonstrated NV head static control, and one demonstrated NV mid-thoracic static control.

**Interpretation:** The results indicate that children with SMA treated with DMTs acquire very little or no unsupported NV head/trunk control. SATCo was shown to be sensitive to differences in trunk control in different types of SMA revealing potential as an outcome measure for therapeutic interventions or treatment effectiveness. Further studies are needed to clarify this potential.

**What this paper adds:**

- Children with SMA1 treated with DMTs do not attain upright head control
- Some children with SMA2/SMA3 treated with DMTs attain upright head/trunk control
- Differences in caudal extent of head/trunk control were found between SMA types
- SATCo is more efficient in identifying upright head/trunk control than functional tests
- Postural control assessment using SATCo has the potential to track treatment outcomes

## Introduction

Spinal muscular atrophy (SMA) is a rare autosomal-recessive disorder caused by mutations in the survival motor neuron 1 (*SMN1*) and characterised by the degeneration of motor neurons of the spinal cord, resulting in muscle weakness and atrophy.^1,2^ The muscles are progressively and symmetrically affected, and weakness is more proximal than distal affecting the lower limbs more than the upper limbs.^1–3^ Although SMA is a single genetically defined disease, those affected range from weak infants to ambulant adults. SMA is classified into phenotypes 0-4 to facilitate management.^4^ This classification is based on the age of onset and highest acquired motor function with SMA type 4 (SMA4) being the less severe form of the disease.^1^ For the three most common types, the functional prospects are limited to ability to move the head and trunk (SMA1), sit (SMA2), and walk (SMA3).^5^

SMA phenotypes, originally determined based on the natural history of the disease, are drastically evolving due to improved standard of care and the introduction of disease-modifying therapies (DMTs).^6^ Three approved DMTs are used in isolation or in combination:^7^ nusinersen (Spinraza®),^8^ onasemnogene abeparvovec (OA, Zolgensma®),^9^ and risdiplam (Evrysdi®).^10^ The survival rate of children with SMA1 treated with DMTs has dramatically increased, with some even attaining independent sitting,^11–13^ one of the main qualifiers of a child with SMA2.

A new major concern is that children with SMA1 able to sit post-DMT are developing spinal deformities at a much younger age, with indications that gained motor functions decline when scoliosis sharply worsens.^14^ The ability to sit, combined with the inability to walk, have long been associated with the development of spinal deformities in SMA.^15^ Children with SMA2 and those with SMA3 who lose ambulation,^16^ have a high risk of developing scoliosis and/or kyphosis.^14,17,18^

The underlying problem is that abnormal trunk postures will eventually develop into fixed spinal deformities. The importance of being able to align the trunk posture with the gravitational vertical (neutral vertical posture, NVP^19^) cannot be understated. However, the ability to achieve head and trunk control in NVP, *per se*, has not been investigated in children with SMA pre-or post-DMT.

The Segmental Assessment of Trunk Control (SATCo) is a validated clinical test^20^ that assesses head and trunk postural control in NV. It identifies the topmost (most cephalo) segment (*head, upper, mid*, and *lower thoracic, upper* and *lower lumbar* spines, or *full trunk*) at which control of the unsupported structure is not demonstrated for each of static, active, and reactive control.

To our knowledge, SATCo has not been previously applied in SMA, but can potentially provide additional information about head and trunk control status in this population, especially relevant with the outstanding advances of DMTs. The first objective of this study was to investigate the extent to which children with various types of SMA treated with DMT can achieve a controlled NVP of the head and trunk. The second objective was to compare these SATCo findings with motor function.

## Method

### Study design, setting, and participants

This was an observational cross-sectional study in a subset of children who participated in the MRC-funded project “Quantification of head and trunk control for children with neuromotor and neuromuscular disorders”, MR/T002034/1. The London-Brent Research Ethics Committee (IRAS project ID: 233469) granted ethical approval, and all parents/guardians were given information about the study before signing a written informed consent.

This convenience subsample of 21 children was screened for eligibility and recruited between 2021-2024 by the clinical staff from four hospitals.

Inclusion criteria were children with SMA between 3 months and 16 years old. Exclusion criteria were both parents/guardians with insufficient understanding of English to give informed consent and children who posed a physical risk during testing to themselves or the research team. Children with fixed spinal deformities and surgical fixation of the spine were included but were SATCo tested only above the deformity or fixation.

In addition to SATCo, data collected from participants included: body height and weight, sex, SMN2 gene copy number, DMT used, age at the start of DMT intake, spine status, and results from functional assessments.

### SATCo assessment protocol

Two trained assessors conducted SATCo. Each child was assessed once for 10-40 min depending on the child’s ability and performance. Three Intel® RealSense™ Depth Cameras D435 were used to record the test (Figure 1).

**Figure 1.**
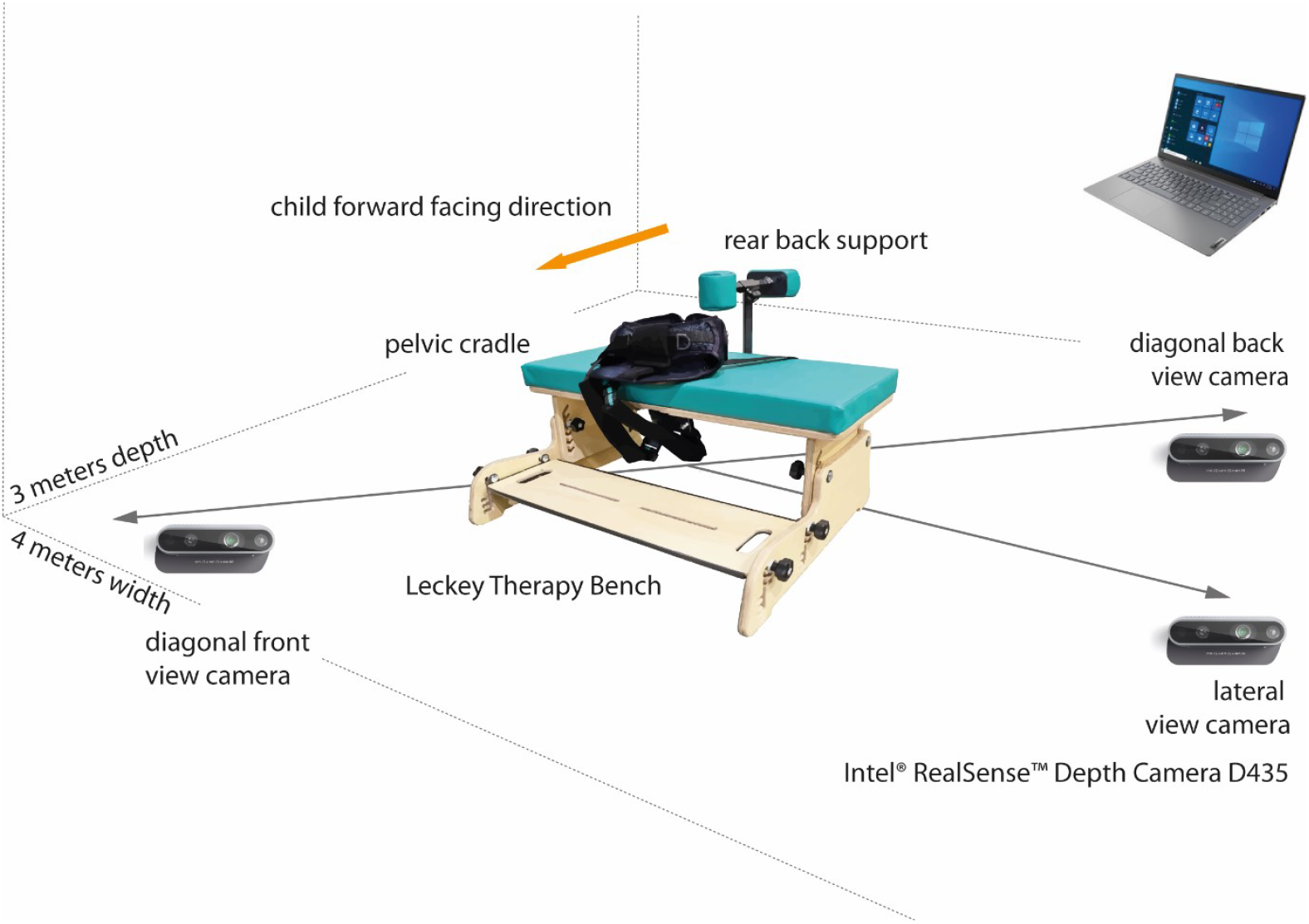
Experimental setup

The child was initially examined to determine the presence of fixed spinal deformities i.e. bony changes that could not be corrected passively. He/she was either (i) positioned prone and asked to rest on the assessor’s hand supporting the sternum 15-30 cm above horizontal or (ii) have his/her spine manually straightened while seated. Once the spine was brought as straight as possible passively, the second assessor manually checked each vertebra from C7 to S1 for fixed/immobile sections. Any spinal deformity was classified as mild, moderate, or severe based on this assessment. This measure was used only to determine the segmental level at which SATCo assessment should stop.

The child was seated on a Leckey Therapy Bench (James Leckey Design Ltd, Ireland) with the pelvis stabilised in NVP and hip and knee flexion at 90 deg. The first assessor conducted the test and checked visually for head/trunk NVP. The second assessor provided a firm horizontal manual support to the trunk and sequentially lowered it to test the segments: H (*head*, support at the shoulder girdle), UT (*upper-thoracic*, axillae), MT (*mid-thoracic*, inferior scapula), LT (*lower-thoracic*, over lower ribs), UL (*upper-lumbar*, below ribs), LL (*lower-lumbar*, pelvis), and FT (*full trunk*, no support given and pelvic support removed).

Sub-components of posture control tested above each supported segment included: *Static*: the ability to maintain for 5 seconds an unsupported head/trunk posture in NV; *Active*: the ability to maintain an unsupported NVP during child-initiated head turn to left and right; *Reactive*: the ability to return to an unsupported NVP following a nudge from the front, back and both sides (excluding the *head* segment).

Each segment and control component was scored as Control Demonstrated, Not Demonstrated (unable to maintain the head/trunk segment above manual support in NVP), or Not Tested (if there was an assessor error).

Conventionally, SATCo is ceased when the child can no longer achieve/maintain an unsupported NVP for any test component.^20^ As this was an exploratory study in a population not previously tested, testing continued to verify the NVP control status of more caudal segments, stopping immediately above any fixed spinal deformities.

Each SATCo was scored by one of three experienced assessors, PBB and TES (authors), and TP, contributor to this study.

## Functional assessments

The functional assessments were conducted within +/-six months of the SATCo, performed by each child’s usual clinical staff (typically physiotherapist). For comparison with SATCo outcomes, items relating to head and trunk control were selected from the World Health Organization Developmental Milestones (WHO),^21^ Hammersmith Infant Neurological Examination (HINE)^22^, Revised Hammersmith Scale (RHS)^23^, and Children’s Hospital of Philadelphia Infant Test of Neuromuscular Disorders (CHOP-INTEND).^24^ Each focuses on different motor functions, and not all are applied in each child.

To find common ground between different functional assessments and SATCo outcomes, we extracted seven motor function categories from all tests performed: *Head control* (while the trunk was supported or not), *Neck flexion while supine, Neck extension while prone, Sitting with support, Sitting independently, Walking with support*, and *Walking independently* (Appendix 1).

This process resulted in two or three items being selected from each functional assessment for each category. To facilitate the determination of attained motor function, the various scores were colour-coded within each item: *unable, partially able*, and *able to perform* (red → yellow → green).

The first category, *Head control while the trunk is supported or not*, was only scored by CHOP-INTEND and HINE. These tests are mostly used for infants and younger children. They relate to the SATCo test of head control, although not specifying the head to be in NVP. No corresponding test was found in RHS or WHO, and older and more capable children were not tested for this category. The only tests related to head control that all children performed were *Neck flexion while supine* and *Neck extension while prone*, with the caveat that a comparison with SATCo is very loose because the child is not upright, and in CHOP-INTEND neck extension is tested *to* the horizontal level and in RHS it is tested *from* the horizontal level upwards.

### Statistical analysis

Statistical analysis was performed using IBM® SPSS® Statistics for Windows version 28.0.1.1, 2022 (IBM Corp., Armonk, NY, USA). SATCo assessment outcomes from 21 children (Control Demonstrated, Control Not Demonstrated, Not Tested) were added to a linear mixed effects (LME) model: factors *condition* (SMA1, SMA2, and SMA3), *segment* (Head, UT, MT, LT, UL, LL, FT), *type of control* (static, active, reactive) were included within fixed effects (grouped by subject), and ANOVA (Satterthwaite approximation) was tested using the SPSS function ‘MIXED’. P≤.05 was considered statistically significant.

## Results

Twenty-one children with various subtypes of SMA were included in this study: twelve with SMA1, seven with SMA2, and two with SMA3 (Table 1) following the inclusion/exclusion criteria. Ten children had mobile spines and 11 presented fixed spinal deformities ranging from mild to severe. All six younger children aged below 3 years had either a straight spine (five children) or mild (one child) spinal deformity. Older children aged between 4-8 years had mild (one child) to severe (five children) spinal deformities (Table 1).

**Table 1.**
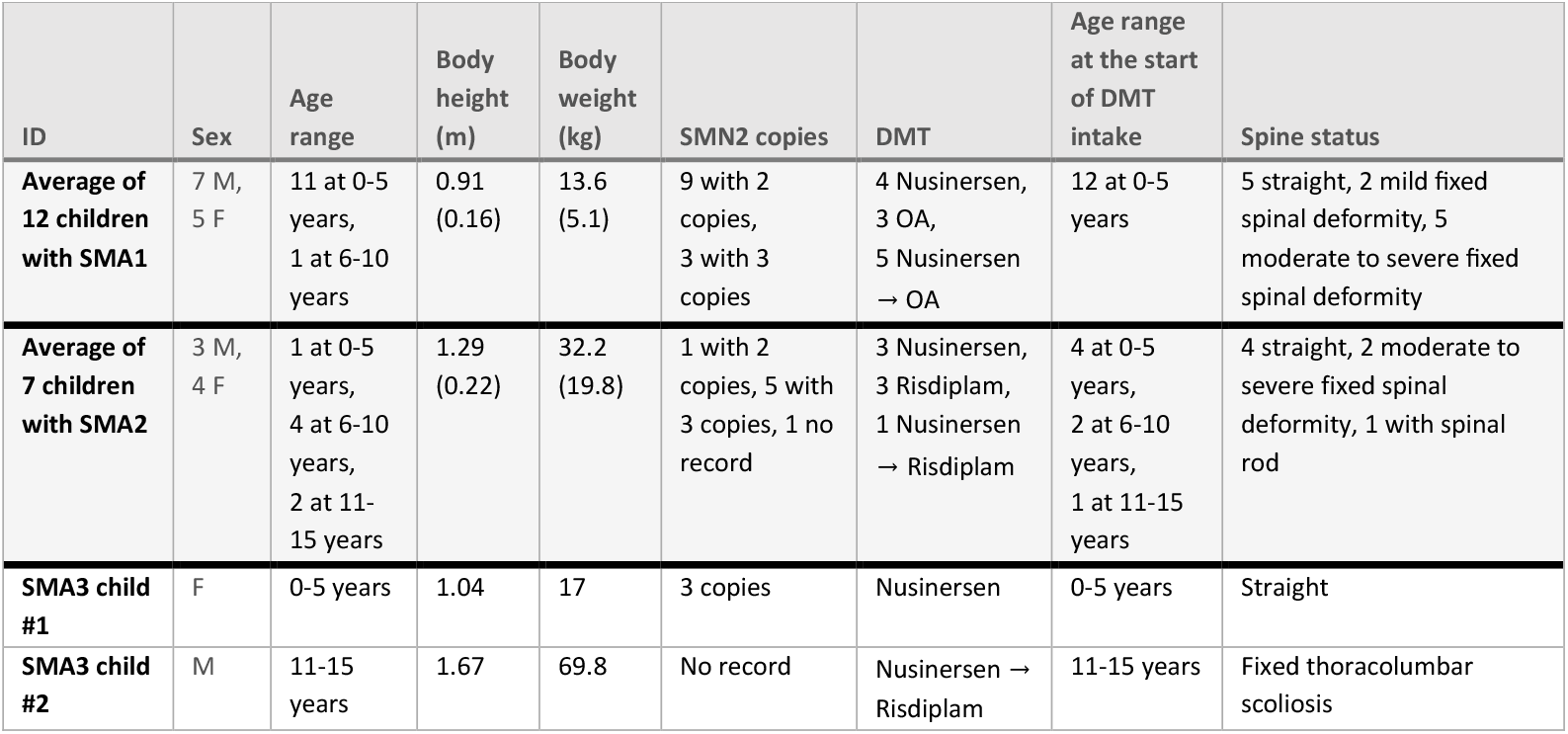
Clinical and anthropometric characteristics of all participants. Body weight and height from from SMA1 and SMA2 were averaged, mean(standard deviation), not from SMA3 because only 2 children were tested. Age and age at start of DMT intake are presented in age ranges: 0-5 years, 6-10 years, 11-15 years.

| ID | Sex | Age range | Body height (m) | Body weight (kg) | SMN2 copies | DMT | Age range at the start of DMT intake | Spine status |
| --- | --- | --- | --- | --- | --- | --- | --- | --- |
| Average of 12 children with SMA1 | 7 M, 5 F | 11 at 0-5 years, 1 at 6-10 years | 0.91 (0.16) | 13.6 (5.1) | 9 with 2 copies, 3 with 3 copies | 4 Nusinersen, 3 OA, 5 Nusinersen → OA | 12 at 0-5 years | 5 straight, 2 mild fixed spinal deformity, 5 moderate to severe fixed spinal deformity |
| Average of 7 children with SMA2 | 3 M, 4 F | 1 at 0-5 years, 4 at 6-10 years, 2 at 11-15 years | 1.29 (0.22) | 32.2 (19.8) | 1 with 2 copies, 5 with 3 copies, 1 no record | 3 Nusinersen, 3 Risdiplam, 1 Nusinersen → Risdiplam | 4 at 0-5 years, 2 at 6-10 years, 1 at 11-15 years | 4 straight, 2 moderate to severe fixed spinal deformity, 1 with spinal rod |
| SMA3 child #1 | F | 0-5 years | 1.04 | 17 | 3 copies | Nusinersen | 0-5 years | Straight |
| SMA3 child #2 | M | 11-15 years | 1.67 | 69.8 | No record | Nusinersen → Risdiplam | 11-15 years | Fixed thoracolumbar scoliosis |

The results of the functional assessments confirmed that motor function in children with SMA1 is generally more compromised than those with SMA2 and SMA3 (Figure 2).

**Figure 2.**
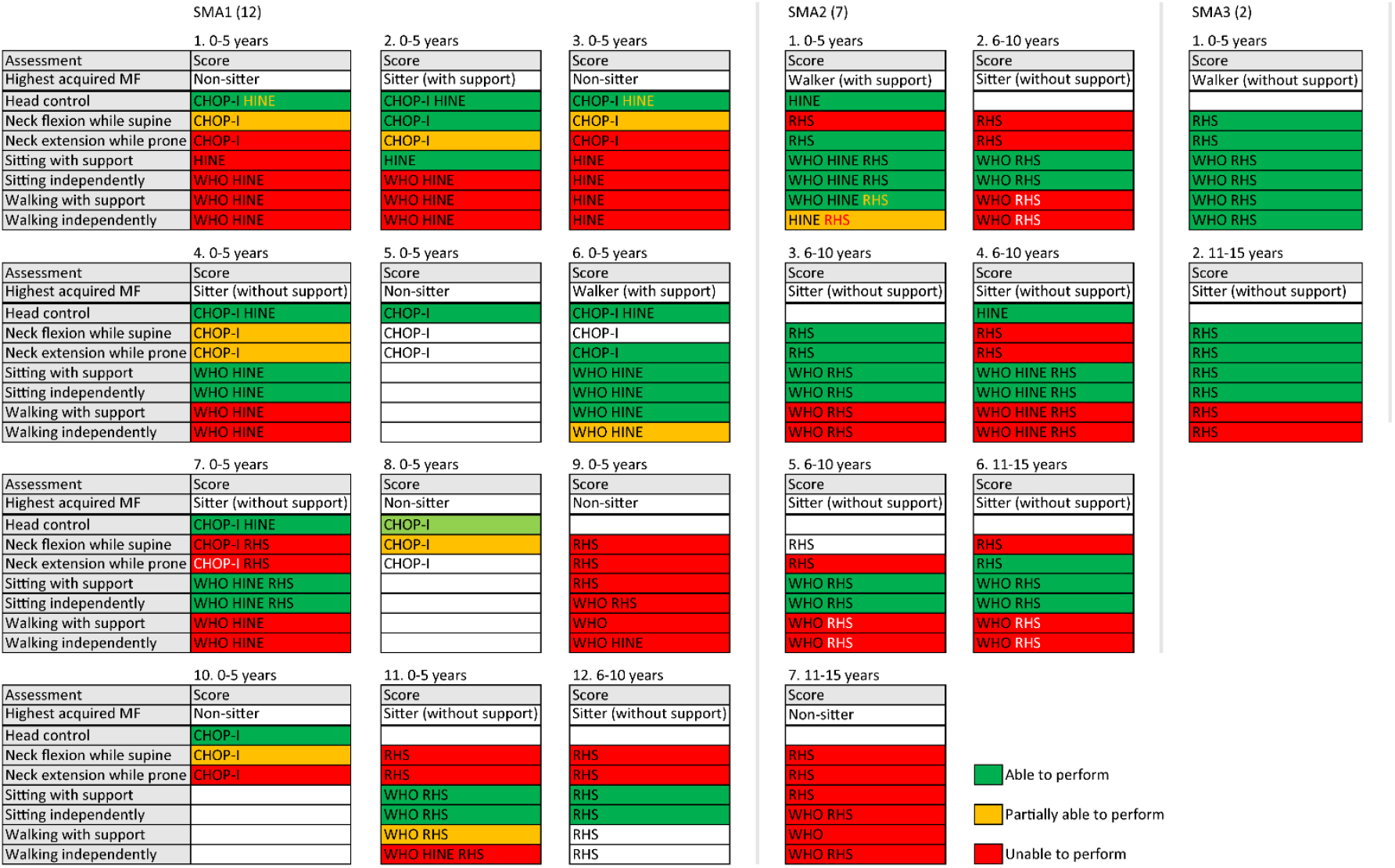
Scores from functional assessments included in SMA REACH UK from all participants. Three columns to the left are from children with SMA1, two columns in the middle, SMA2, and one column to the right, SMA3. Participants are organised by age range, starting from the top-left.

Of the 12 children with SMA1, nine were tested for head control (CHOP-INTEND item #12 and/or HINE item #1), and eight were identified as having functional control (88.9%). Most were unable to flex or extend their necks while horizontal (8.3%). Only two children with SMA2 were tested for and demonstrated head control (100%), and no child with SMA3 was tested for head control. Three out of seven children with SMA2 were able to extend their necks (42.9%), and one of them was also able to flex his neck (14.3%). Both children with SMA3 fully scored in these categories (100%).

Five children with SMA1 could sit independently (41.7%), and one could also walk with support (8.3%). In contrast, six children with SMA2 could sit independently (85.7%), and one could also walk with support (14.3%). Both children with SMA3 could sit independently (100%), and one walked independently (50%).

SATCo results revealed that none of the children with SMA1 demonstrated NV head control or any NV trunk control. Three of the seven children with SMA2 and one with SMA3 demonstrated static NV head control. One of the three children with SMA2 who demonstrated NV head control also demonstrated upper- and mid-thoracic static NV trunk control. No child demonstrated active or reactive NV control at any segment (Figure 3).

**Figure 3.**
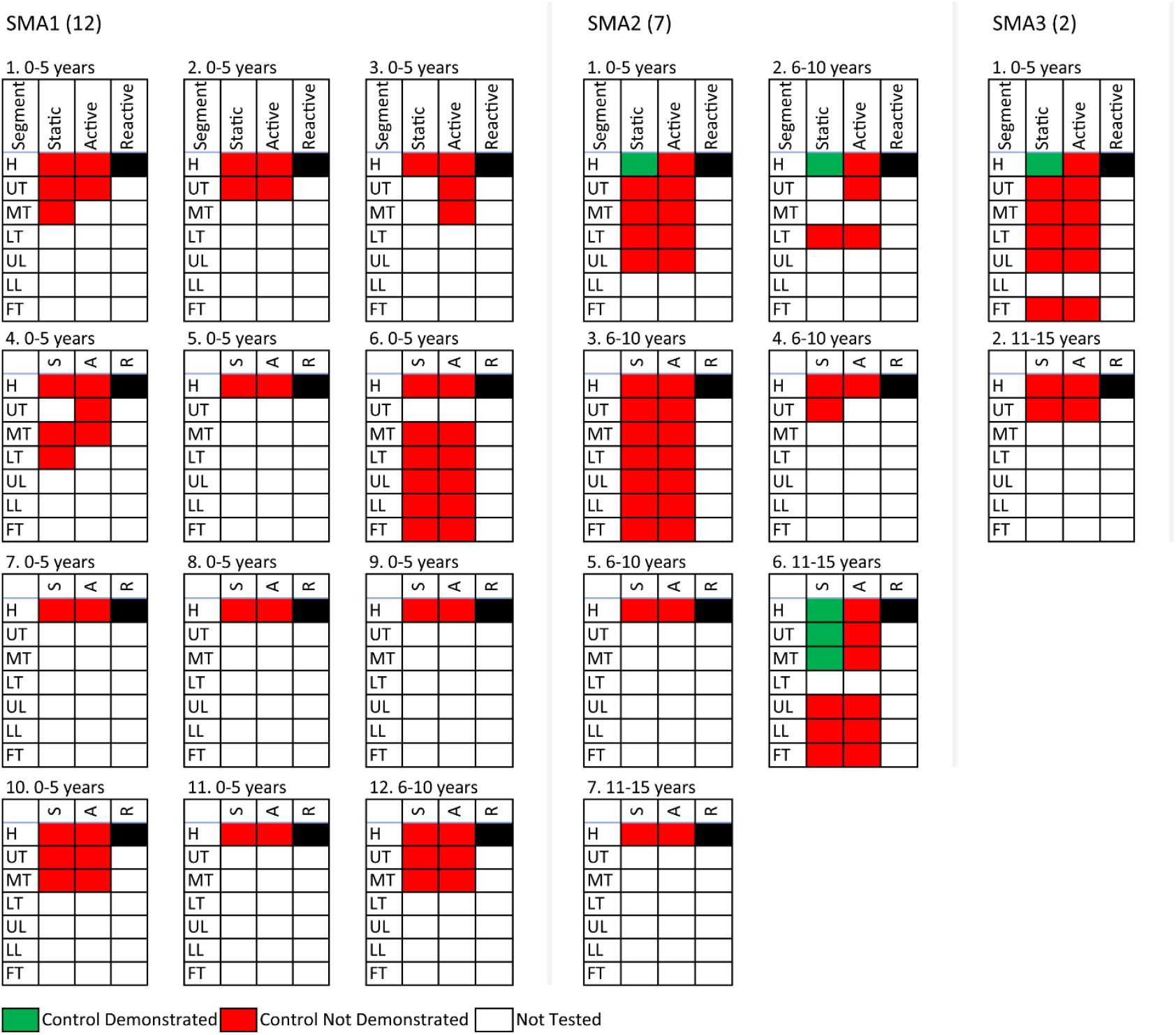
SATCo scores from all participants. Three columns to the left are from children with SMA1, two columns in the middle, SMA2, and one column to the right, SMA3. Participants are organised by age range, starting from top-left.

The results from LME analysis showed differences in SATCo scores related to the *condition* (F=4.96, p<.05) and *control type* being tested (F=6.40, p<.05), not the *segment* (F=1.26, p=.28). The significant interaction *condition* * *control type* (F=3.14, p<.05) shows that difference in results found between SMA types was related to the control type.

These results present a marked contrast to the outcomes of the tests of motor function. From the four children who demonstrated any extent of NVP control (three SMA2 and one SMA3), only one was tested for and demonstrated head control with HINE. Three demonstrated neck extension while prone but only the child with SMA3 demonstrated neck flexion while horizontal. All four children showed functional independent sitting ability. Both children with SMA2 and SMA3 who could walk, with or without support, demonstrated static NV head control. The one child with SMA2 who scored MT static NV static control was unable to walk (Figure 4). She was also the oldest with no fixed spinal deformities and could sit independently.

**Figure 4.**
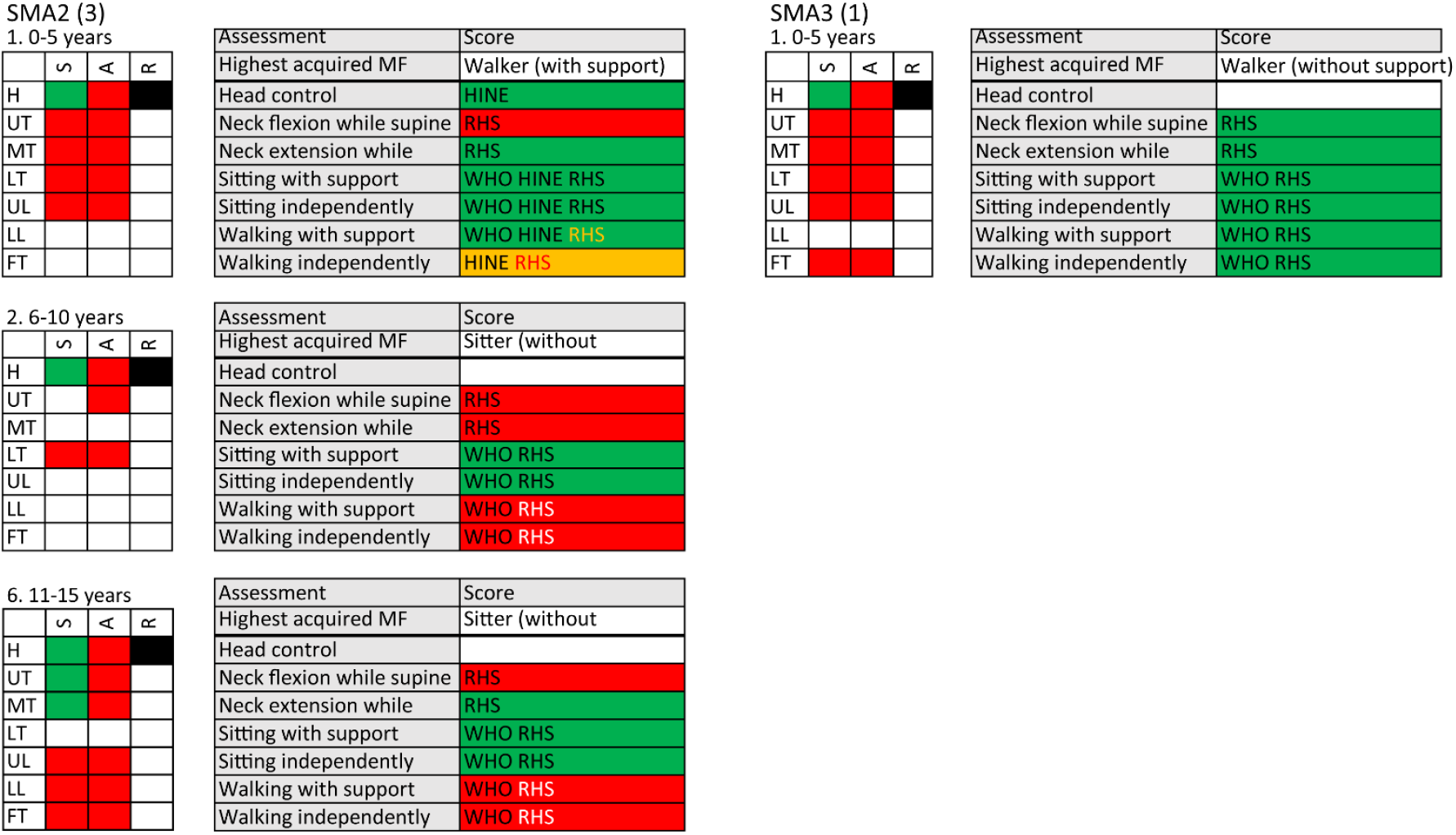
SATCo and functional assessment scores from four participants who demonstrated some degree of NVP control. Left columns are from children with SMA2, and right columns are from a child with SMA3.

Regarding DMT type, the children with SMA2 and SMA3 who demonstrated any NVP control were treated with Risdiplam (1) and Nusinersen (3). However, other children with the same phenotypes were also treated with these drugs and did not obtain NVP control.

SATCo outcomes were very consistent overall. However, the sample size was small and to perform any statistical correlation with functional assessment scores, DMT type, age at start of DMT intake, and SMN2 gene copy number, was not appropriate.

## Discussion

SATCo was used to assess NVP segmental head/trunk control in 21 children with three subtypes of SMA under treatment with three types of DMTs. A significant difference was found between groups. No child at the more severe end of the disease spectrum (SMA1) demonstrated NVP head control. Three children with SMA2 and one with SMA3 demonstrated NV static head control. However, none of the children tested demonstrated active or reactive NVP control.

### Functional tests and SATCo: what do they assess

Functional tests are designed to demonstrate and classify the achievement of everyday functional abilities. These tests are sometimes additionally used to infer head and trunk control status within the postures used. In general, functional assessments impose few limitations on head/trunk postures or strategies used. As an example, sitting is often described as ‘upright’ for full credit, but with no definition of upright posture.

In contrast, SATCo has the specific objective of determining the control status of the unsupported column of segments above the manual support in NVP. NVP has a precise definition^19^ with the critical element that no joint should be at its end range, specifically within the spine. Clearly, fixed spinal deformities do not match this definition, moving the focus to testing trunk control only above the start of the fixed curve. This strict definition of NVP means that movement control in this posture i.e. the ability to control the magnitude of the moment arms/moments at various joints, is assessed in all directions of movement available at the segment concerned (a segment comprising a small number of joints and associated structures). SATCo assesses the underlying control mechanisms that bring about movement and function. It complements and clarifies the results of functional tests by providing this fundamental information. Figure 5 provides an illustration of these principles.

**Figure 5.**
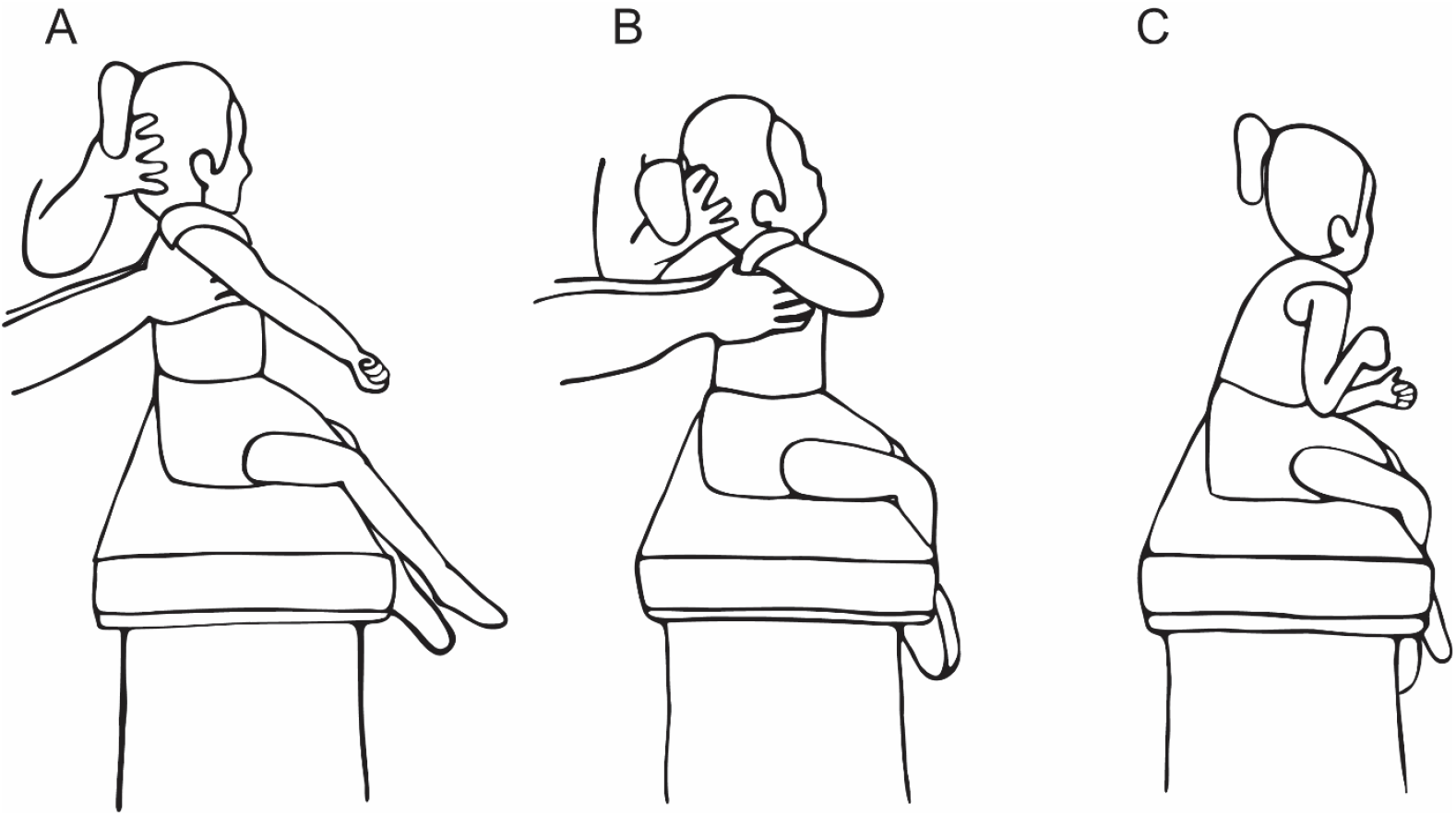
SATCo test of NV head control in a child with SMA1 (lateral view). The child’s pelvis was supported by a harness secured to the bench. (A) Child held with the upper thoracic spine in NV as required by SATCo in order to test head control (cervical spine control): unable to hold her head upright and requiring constant head support. (B) Child held with the trunk less upright: briefly able to bring her head near upright. (C) Child in her preferred sitting posture with lumbar and thoracic spine collapsed into flexion: able to maintain her head upright for longer intervals by resting in full cervical extension with lateral head support provided by the shoulder girdle. This posture could be held without active control.

### The clinical value of SATCo in children with SMA

This study has shown that, for children with SMA1, DMTs alone do not lead to head and trunk NVP control. Sitting is a meaningful achievement enabled by DMTs but control progression, leading to greater functional ability, is a head-downwards process. The next step is to investigate means to improve control acquisition and thus ability to sit aligned with the gravitational vertical.

SATCo revealed that NVP control acquisition was greater in SMA2 and SMA3. A recent study showed a reduced rate of progression of scoliosis in children with SMA2 who had early DMT treatment,^25^ suggesting a greater ability to control NVP. The only child with SMA2 who demonstrated mid-thoracic static NVP control from our cohort was the oldest who had no fixed spinal deformities. This supports future exploratory work.

The progression of muscle weakness is continuous in all SMA types throughout their lifespan.^15^ The management of children with different types of SMA might benefit from the use of SATCo to determine the core head/trunk control status for a more complete and systematic picture. Targeted Training (TT), the therapy strategy linked with SATCo, helps a child gain head/trunk NVP control.^26–29^ Once SATCo determines at which segment control is no longer demonstrated, the TT equipment is adjusted so that his/her body is firmly supported in NV below this targeted segment, and the child is encouraged to maintain all segments above the support in NVP. Once the child achieves this NVP control, the firm support is lowered for him/her to start training the next caudally-directed segment, reducing the control learning load to manageable proportions. TT could potentially address the dichotomy between allowing free trunk movement to strengthen muscles or using a rigid trunk bracing to reduce fatigue or minimise development of fixed spinal deformities.^30^ At this stage, it is unknown whether these children have not achieved NV head/trunk control due to lack of efficacy of DMTs or due to lack of access to TT type programs amongst this patient group.

### Limitations

This was a small exploratory study, with a small sample size. The process of enabling the outcomes of functional tests to be viewed in a manner that could be directly compared to SATCo outcomes proved challenging and different methods may emerge in the future. This also meant that a direct comparison of functional attainment and head/trunk control as assessed by SATCo was not possible.

### Conclusion and future studies

The introduction of DMTs represent a breakthrough in the treatment of children with SMA. Many children with SMA1 post-DMT are surviving and can sit, but the results from this study indicate that they do not acquire unsupported NVP head/trunk control, which is critical for functional gains. A small number of children with SMA2 and SMA3 attained NVP static head control, and the commencement of static trunk control, but none demonstrated any active or reactive control.

SATCo results are noticeably different from the results from functional assessments due to the different basis of the tests. The focus on function means that assessments do not need to address strategies the children might use to accomplish these activities, such as arm support. SATCo highlights the deficits in unsupported NVP head/trunk control in SMA that form the basis for function to emerge and evolve. The problem of developing fixed spinal deformities by sustaining a collapsed sitting posture for long periods has been a constant concern for children with SMA2 but is a new concern for children with SMA1 post-DMT. The extent to which DMTs can reduce this risk when combined with (physio)therapy treatments such as Targeted Training, which focuses on NV head/trunk posture control, is currently unknown and should be investigated in future longitudinal studies and randomized clinical trials.

## Data Availability

The data that support the findings of this study will be available on request from the corresponding author in the near future. The data are not publicly available due to privacy or ethical restrictions.

## Abbreviations

CHOP-INTEND: CHOP-I The Children’s Hospital of Philadelphia Infant Test of Neuromuscular Disorders
HINE: Hammersmith Infant Neurological Examination
LME: Linear mixed-effects model
NV NVP: Neutral vertical, neutral vertical posture
OA: Onasemnogene abeparvovec
RHS: Revised Hammersmith Scale
SATCo: Segmental Assessment of Trunk Control
SMA: Spinal muscular atrophy
SMA0, SMA1, SMA2, SMA3, SMA4: Spinal muscular atrophy types 0, 1, 2, 3, and 4
SMN: Survival motor neuron
WHO: World Health Organization

## Acknowledgments

We gratefully acknowledge the contribution to this study made by Teresa Perez (TP) and Natalia Rajkowska from Manchester Metropolitan University. We also thank James Leckey Design Ltd (Lisburn, Ireland) for kindly providing Leckey Therapy Benches for this study.

## Supporting Information

**Appendix 1.**
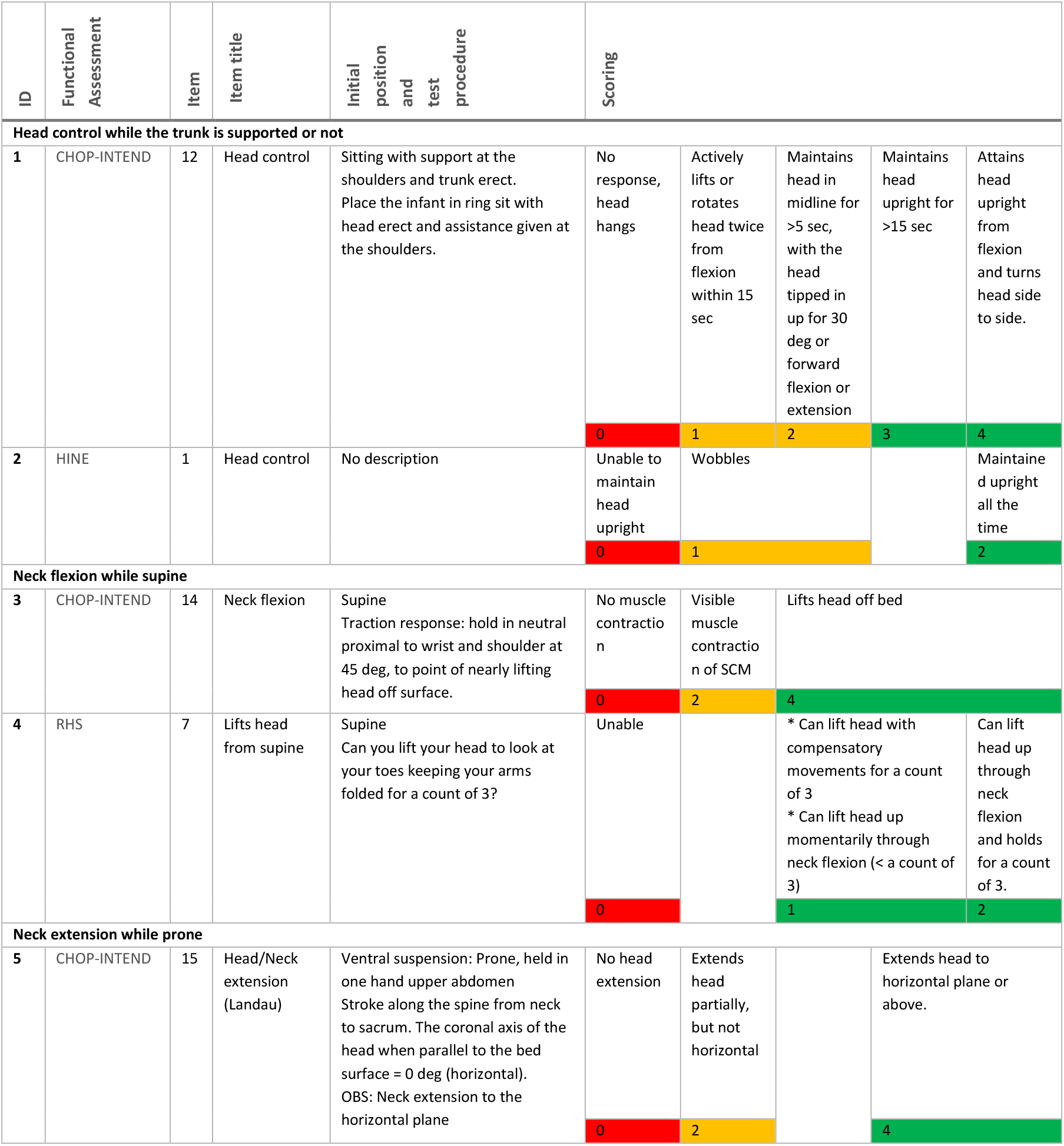
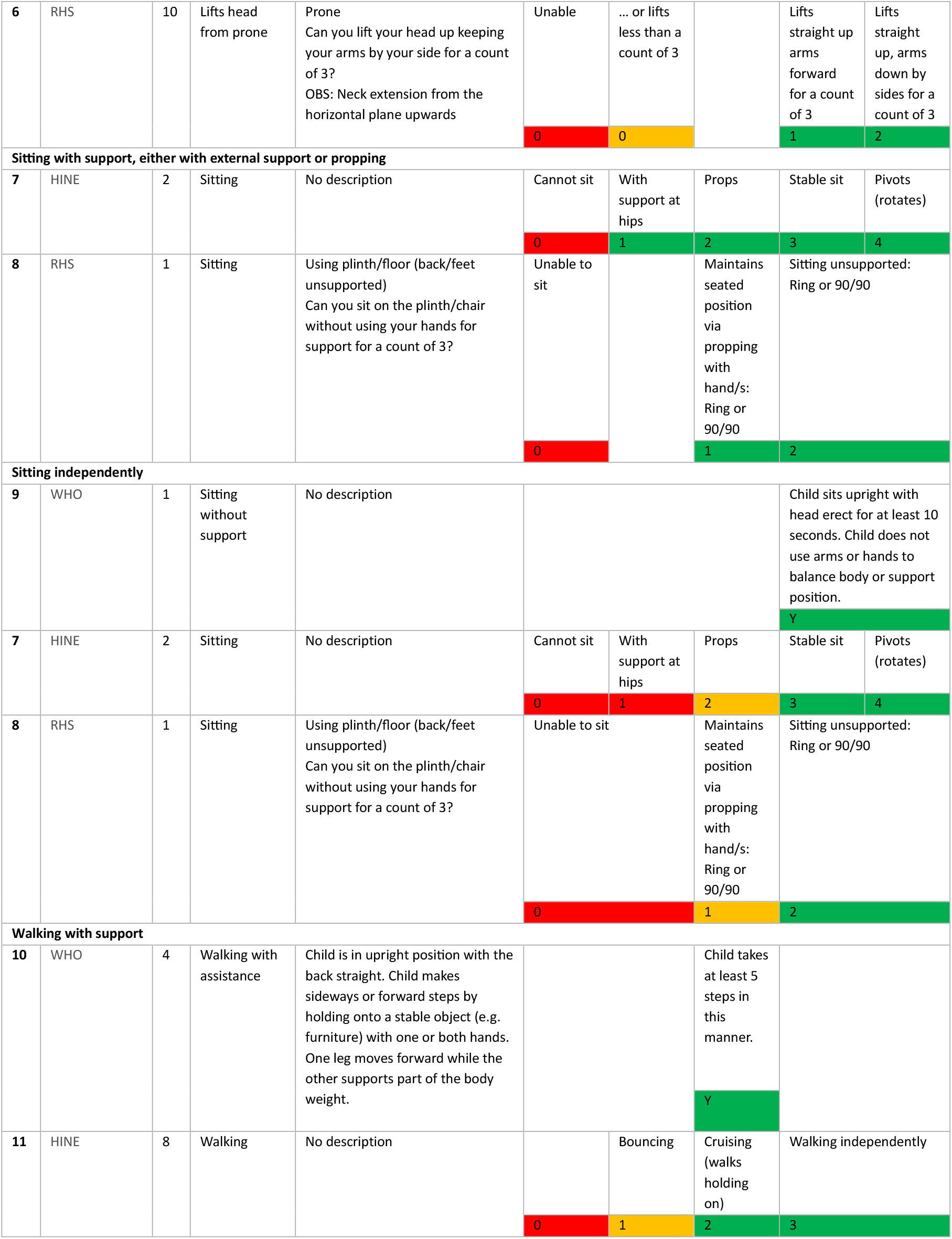
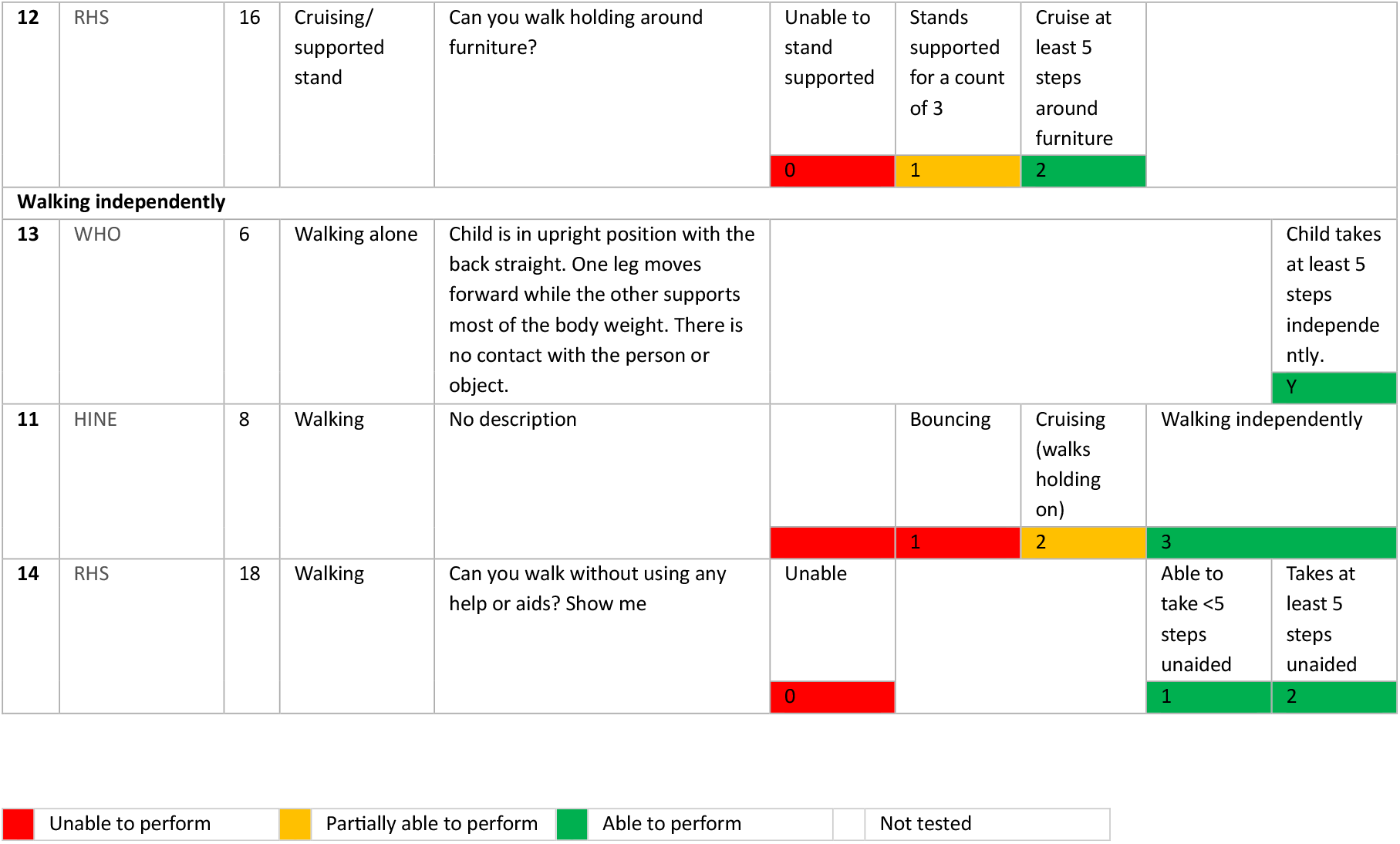
Seven categories of motor function and corresponding items selected from functional assessments performed in children with SMA. For each category, items from 2-3 tests were grouped and color-coded into 2-3 levels: unable, partially able, and able to perform (red → yellow → green). Categories not tested are left blank. Each item’s number in its corresponding assessment is shown in the column “Item”, and all descriptions about the test procedure are shown in the column “Initial position and test procedure”.

## References

1. Munsat TL, Davies KE. International SMA consortium meeting (26–28 June 1992, Bonn, Germany). Neuromuscular Disorders 1992; 2: 423–8.

2. Prior TW, Leach ME, Finanger E. Spinal Muscular Atrophy. In: GeneReviews®, December 3, 2020. Seattle, USA: University of Washington, 1993. https://www.ncbi.nlm.nih.gov/books/NBK1352/ (accessed 30 July 2024).

3. Carter GT, Abresch RT, Fowler WMJ, Johnson ER, Kilmer DD, McDonald CM. Profiles of neuromuscular diseases: Spinal muscular atrophy. Am J Phys Med Rehabil 1995; 5: S150–9.

4. Lunn MR, Wang CH. Spinal muscular atrophy. Lancet 2008; 371: 2120–33.

5. Lin CW, Kalb SJ, Yeh WS. Delay in diagnosis of spinal muscular atrophy: A systematic literature review. Pediatr Neurol. 2015; 53: 293–300.

6. Tizzano EF, Finkel RS. Spinal muscular atrophy: A changing phenotype beyond the clinical trials. Neuromuscular Disorders 2017; 27: 883–9.

7. Proud CM, Mercuri E, Finkel RS, et al. Combination disease-modifying treatment in spinal muscular atrophy: A proposed classification. Ann Clin Transl Neurol 2023; 10: 2155–60.

8. Finkel RS, Mercuri E, Darras BT, et al. Nusinersen versus Sham Control in Infantile-Onset Spinal Muscular Atrophy. New England Journal of Medicine 2017; 377: 1723–32.

9. Mendell JR, Al-Zaidy S, Shell R, et al. Single-Dose Gene-Replacement Therapy for Spinal Muscular Atrophy. New England Journal of Medicine 2017; 377: 1713–22.

10. Baranello G, Darras BT, Day JW, et al. Risdiplam in Type 1 Spinal Muscular Atrophy. New England Journal of Medicine 2021; 384: 915–23.

11. Mercuri E, Sumner CJ, Muntoni F, Darras BT, Finkel RS. Spinal muscular atrophy. Nat Rev Dis Primers 2022; 8. DOI:10.1038/s41572-022-00380-8.

12. Antonaci L, Pera MC, Mercuri E. New therapies for spinal muscular atrophy: where we stand and what is next. Eur J Pediatr 2023; 182: 2935–42.

13. Aragon-Gawinska K, Daron A, Ulinici A, et al. Sitting in patients with spinal muscular atrophy type 1 treated with nusinersen. Dev Med Child Neurol 2020; 62: 310–4.

14. Al Amrani F, Amin R, Chiang J, et al. Scoliosis in Spinal Muscular Atrophy Type 1 in the Nusinersen Era. Neurol Clin Pract 2022; 12: 279–87.

15. Wadman RI, Wijngaarde CA, Stam M, et al. Muscle strength and motor function throughout life in a cross-sectional cohort of 180 patients with spinal muscular atrophy types 1c–4. Eur J Neurol 2018; 25: 512–8.

16. Fujak A, Raab W, Schuh A, Richter S, Forst R, Forst J. Natural course of scoliosis in proximal spinal muscular atrophy type II and IIIa: Descriptive clinical study with retrospective data collection of 126 patients. BMC Musculoskelet Disord 2013; 14. DOI:10.1186/1471-2474-14-283.

17. Wishart BD, Kivlehan E. Neuromuscular scoliosis: When, who, why and outcomes. Phys Med Rehabil Clin N Am 2021; 32: 547–56.

18. Wijngaarde CA, Brink RC, De Kort FAS, et al. Natural course of scoliosis and lifetime risk of scoliosis surgery in spinal muscular atrophy. Neurology 2019; 93: E149–58.

19. Butler PB, Major R. Segmental Assessment of Trunk Control (SATCo). https://optimi.org.uk/satco/ (accessed 19 Sept 2024).

20. Butler PB, Saavedra S, Sofranac M, Jarvis SE, Woollacott MH. Refinement, reliability, and validity of the segmental assessment of trunk control. Pediatric Physical Therapy 2010; 22: 246–57.

21. De Onis M. WHO Motor Development Study: Windows of achievement for six gross motor development milestones. Acta Paediatrica, International Journal of Paediatrics 2006; 95: 86– 95.

22. Bishop KM, Montes J, Finkel RS. Motor milestone assessment of infants with spinal muscular atrophy using the hammersmith infant neurological Exam—Part 2: Experience from a nusinersen clinical study. Muscle Nerve 2018; 57: 142–6.

23. Ramsey D, Scoto M, Mayhew A, et al. Revised Hammersmith Scale for spinal muscular atrophy: A SMA specific clinical outcome assessment tool. PLoS One 2017; 12. DOI:10.1371/journal.pone.0172346.

24. Glanzman AM, Mazzone E, Main M, et al. The Children’s Hospital of Philadelphia Infant Test of Neuromuscular Disorders (CHOP INTEND): Test development and reliability. Neuromuscular Disorders 2010; 20: 155–61.

25. Coratti G, Lenkowicz J, Pera MC, et al. Early treatment of type II SMA slows rate of progression of scoliosis. J Neurol Neurosurg Psychiatry 2023; 95: 235–40.

26. Butler PB. A preliminary report on the effectiveness of trunk targeting in achieving independent sitting balance in children with cerebral palsy. Clin Rehabil 1998; 12: 281–93.

27. Pin TW, Butler PB, Shum SLF. Targeted Training in managing children with poor trunk control: 4 case reports. Pediatric Physical Therapy 2018; 30: E8–13.

28. Butler PB, Major RE. The learning of motor control: Biomechanical considerations. Physiotherapy 1992; 78: 6–11.

29. Curtis DJ, Holbrook P, Bew S, Ford L, Butler P. Functional change in children with cerebral palsy. ArXiv e-prints 2018; : 1–14.

30. Peeters LHC, Janssen MMHP, Kingma I, van Dieën JH, de Groot IJM. Patients With Spinal Muscular Atrophy Use High Percentages of Trunk Muscle Capacity to Perform Seated Tasks. Am J Phys Med Rehabil 2019; 98: 1110–7.

